# Characterization of baseline and longitudinal DNA Methylation in patients with sporadic Parkinson’s disease

**DOI:** 10.1101/2023.09.28.23296098

**Authors:** Paulina Gonzalez-Latapi, Bernabe Bustos, Siyuan Dong, Steven Lubbe, Tanya Simuni, Dimitri Krainc

## Abstract

**Objective:** To characterize DNA methylation differences between sporadic Parkinson’s Disease and healthy control individuals enrolled in the Parkinson’s Progression Markers Initiative.

**Methods:** We characterized cross-sectional and longitudinal DNA methylation differences between individuals with sporadic (i.e., non-genetic) PD and healthy controls. We included 282 individuals (196 Parkinson’s Disease individuals and 86 healthy control individuals). DNA methylation data was collected at the time of enrollment and longitudinally over three years.

**Results:** This analysis revealed 81,604 differentially methylated positions and 5,281 differentially methylated regions between sporadic PD and healthy controls. Gene ontology analysis revealed that these differentially methylated positions and regions were associated with genes involved in diverse cellular processes, including several with specific functions in the brain (Focal adhesion”, “Cholinergic synapse”, “Glutamatergic synapse”, “Dopaminergic synapse”). Integration of both differentially methylated sites and expressed genes showed 20 genes that were hypomethylated and overexpressed and one gene, CTSH that was hypermethylated and associated with reduced expression.

**Interpretation of Results:** Our study provides evidence that alterations in the methylome in Parkinson’s Disease are discernible in blood, evolve over time, and reflect cellular processes linked to ongoing neurodegeneration. These findings lend support to the potential of blood DNA methylation as an epigenetic biomarker for Parkinson’s Disease. To fully comprehend DNA methylation changes throughout the progression of Parkinson’s Disease, additional profiling at longer intervals and during the prodromal stage will be necessary.

## Introduction

Parkinson’s Disease (PD) is the second most common neurodegenerative disorder, and its prevalence is expected to double by 2040 ^1, 2^. Although various genetic causes of PD have been identified, genetic factors account for only a minority of disease heritability ^3^. Accumulating epidemiological evidence has shown that environmental factors can increase the risk of PD ^4^, but not everyone responds in the same way to environment. Presently, it appears that the complex interplay between genetic and environmental factors underlies the pathogenesis of PD ^5^. Some of the features of PD risk may be reflected in the epigenome, which serves as an interface between genetic and environmental risk factors.

Epigenetic modifications do not affect the DNA sequence and can result in alterations in gene expression. DNA methylation (DNAm) is the most widely studied epigenetic phenomenon ^6-8^. As such, there is growing interest in exploring the role of DNAm in PD. Some studies suggest that methylation patterns in the blood of PD patients differ between cases and controls and may be related to the onset of PD ^8-10^. However, most of these studies have been conducted in heterogeneous or small cohorts. Therefore, there is a need for large-scale studies involving DNA samples from deeply clinically and biologically characterized PD cases and suitable controls, with rigorous collection and storage protocols.

The Parkinson’s Progression Markers Initiative (PPMI) is an observational, international, multicenter study designed to establish biomarker-defined cohorts and identify clinical, imaging, genetic, and biospecimen (e.g., blood sample, cerebrospinal fluid) markers of PD progression. PPMI was established in 2010, and participants have been followed longitudinally, with clinical and biological samples, including DNAm, collected on a yearly basis ^11^ Longitudinal studies with multiple time points where methylation data was assessed, could provide novel insights into methylation trajectories in disease ^12^.

The aim of this study was to characterize DNAm differences between individuals with sporadic PD (i.e. non-genetic) and healthy controls (HC) enrolled in the PPMI study. Importantly, we report on cross-sectional analyses as well as longitudinal DNAm changes in these individuals. Additionally, we provide initial data on the use of DNAm to develop an epigenetic signature to differentiate sPD and HC individuals.

## Methods

### Study cohort

Data were downloaded from PPMI LONI database (December 13, 2021). The aims and methodology of PPMI have been published ^11^. For this study, we used the analytical dataset for sporadic Parkinson Disease (sPD) and HC participants. All study participants were enrolled at the initial stage of PPMI. Importantly, sPD individuals were treatment naïve and within two years of diagnosis at enrollment. Individuals with a variant in a known PD gene were excluded from this analysis. Individuals with a known condition that could impact DNAm (i.e. diagnosis of a blood or bleeding disorders, past medical history of cancer, autoimmune conditions and inflammtory bowel disease) were also excluded from our analysis.

### DNA methylation analysis

For each study participant, DNAm data of ∼ 850,000 CpG-sites were obtained from the Infinium MethylationEPIC array (Illumina) generated using bisulfite converted blood DNA. Raw IDAT files were available from PPMI. The *ChAMP* pipeline ^13, 14^ for Infinium MethylationEPIC BeadChip (Illumina) was used for all analyses, following the default workflow. Raw IDAT files were exported for processing in R (version 4.1). While β values were used for interpretation of the results, *M* values [*M* value□= □log2(methylated intensity/unmethylated intensity)] were used for the differential methylation analysis ^15^.Quality control and filtering steps were applied to exclude probes with detection *p-*value > 0.01, probes with < 3 beads in at least 5% of samples, non-CpG probes, probes that fall in single-nucleotide polymorphism (SNP), multi-hit probes and probes located in sex chromosomes ^16^. Data were normalized with BMIQ Method ^17^ and batch effects were corrected by *ComBat* ^18^. Cell type influence on the whole blood data was corrected with *RefbaseEWAS* ^19^. Differentially methylated positions (DMPs) were detected using limma ^20^. For the longitudinal analysis, we fit linear regression models with cluster-robust standard errors using the robust_se( ) function in the R sandwich package. Age, disease duration, sex, blood cell composition and levodopa equivalent daily dose (LEDD) were included as covariates in these analyses.

Differentially methylated regions (DMRs) were detected using the *DMRcate* ^21^. A bandwidth of 1000 nucleotides (lambda□=□1000) and a scaling factor of 2 (*C*□=□2) were used as recommended by the DMRcate authors. An adjusted (FDR) *p* value <0.05 was considered statistically significant for all methylation analyses.

We will have over 80% power at 70% of methylation sites to detect a difference in methylation at significance level 9e-8, as recommended for the EPIC array ^22^.

### RNASeq analyses

Differential gene expression analyses were performed only for the genes that were mapped to or near (± 1000 base pairs) to the DMRs. Blood gene expression was obtained from the PPMI RNASeq raw read counts. Differential expression analysis was done using the R package *DESeq2* ^23^, and an adjusted FDR *p* value <0.05 was used to select significantly differentially expressed genes.

## Results

Our study included a total of 282 participants from the PPMI cohort. Among these, 196 individuals belonged to the sPD group, with 34% being female, mean age of 63.31 years, and a mean disease duration from diagnosis of 0.5 years at enrollment. Additionally, 86 healthy control participants were included, with 32% being female and a mean age of 63.95 at enrollment. There were no significant differences in age at DNA collection, sex ratio, or racial composition between the sPD and healthy control groups **(Table 1)**.

**Table 1.** Demographic characteristics at baseline.

|  | **sPD(n=196)** | **HC (n=86)** | ***p* (95% CI)** |
| --- | --- | --- | --- |
| Mean age at baseline visit (SD) | 61.80 (9.36) | 62.15 (10.78) | 0.795 (-2.30,2.99) |
| Female/Male (n) | 58/129 | 28/58 | 0.79 |
| White (n) | 185 | 77 | 0.38 |
| Median disease duration at baseline (range) | 0.4 (0 -2) | NA | NA |
| Median symptom age (range) | 62 (35-81) | NA | NA |

Baseline and longitudinal clinical characteristics are presented in **Table 2**. Individuals with sPD exhibited significantly lower Montreal Cognitive Assessment (MoCA) scores compared to HC, although their scores still fell within the range for normal cognition. As expected, both motor and non-motor scores were significantly worse in sPD individuals, and these scores declined further during follow-up. None of the sPD participants were taking any dopaminergic medication at baseline.

**Table 2.**
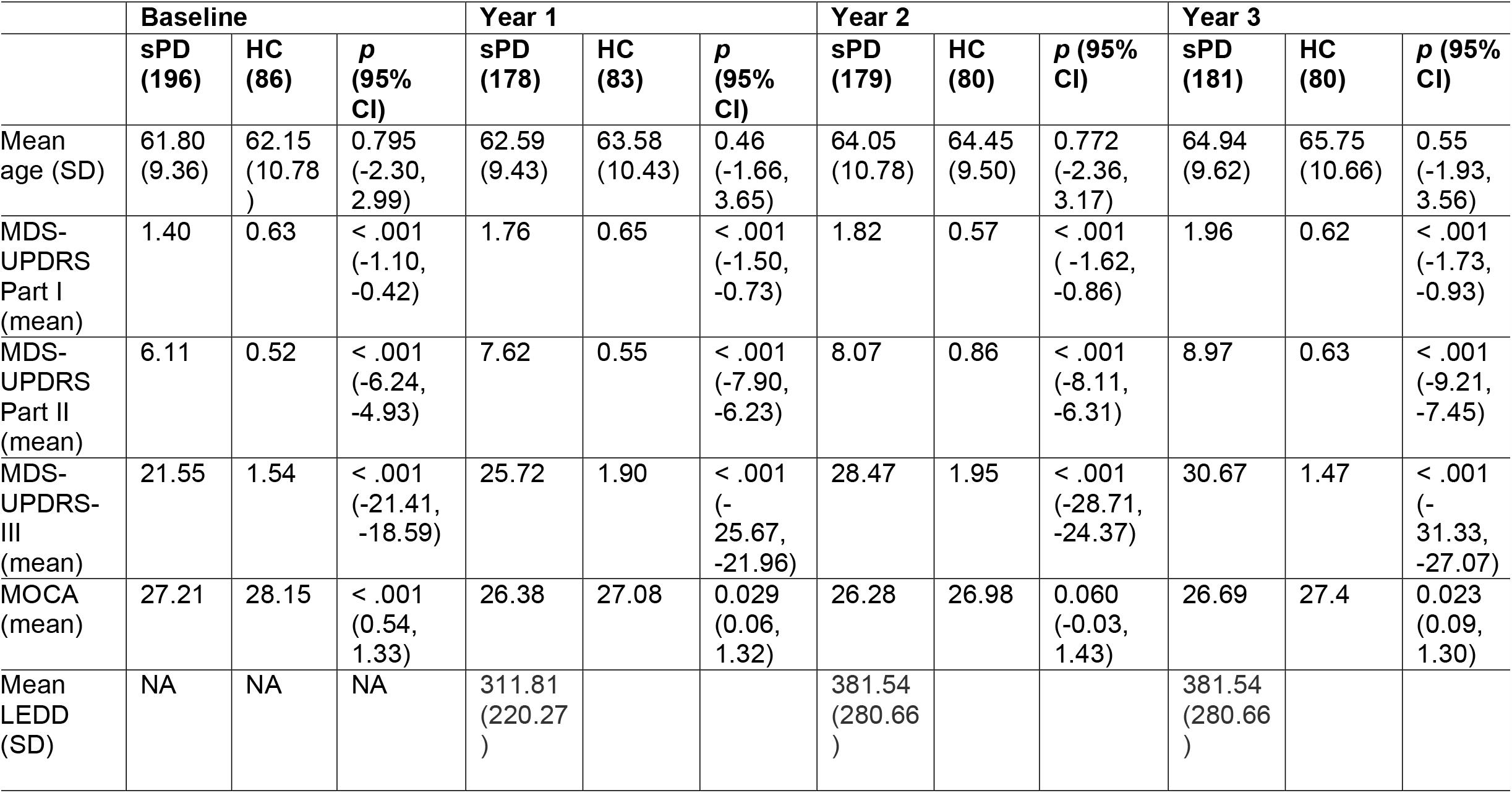
Clinical characteristics at baseline and on longitudinal follow up.

### Blood cell composition differs between sPD and healthy controls

Given that we used whole blood DNA for methylation profiling, we considered the potential impact of differential lymphocyte cell type distributions on our analysis. To address this, we utilized distinctive cell-specific methylation profiles to estimate the proportional abundance of blood cell types in our samples based on the EPIC array’s specific probes. At baseline, the sPD group exhibited a significantly higher proportion of granulocytes compared to HC (*t test:* 0.64 vs. 0.62, p < 0.0001), as shown in **Fig1**. This difference persisted during follow-up. Additionally, at 3-year follow up the proportion of CD8+ and CD4+ cells decreased in the sPD group and was significantly lower compared to the HC group (0.051 vs 0.060, p=0.041 for CD8+; 0.127 vs 0.148, p = 0.0001 for CD4+) (**Fig1**).

**Figure 1.**
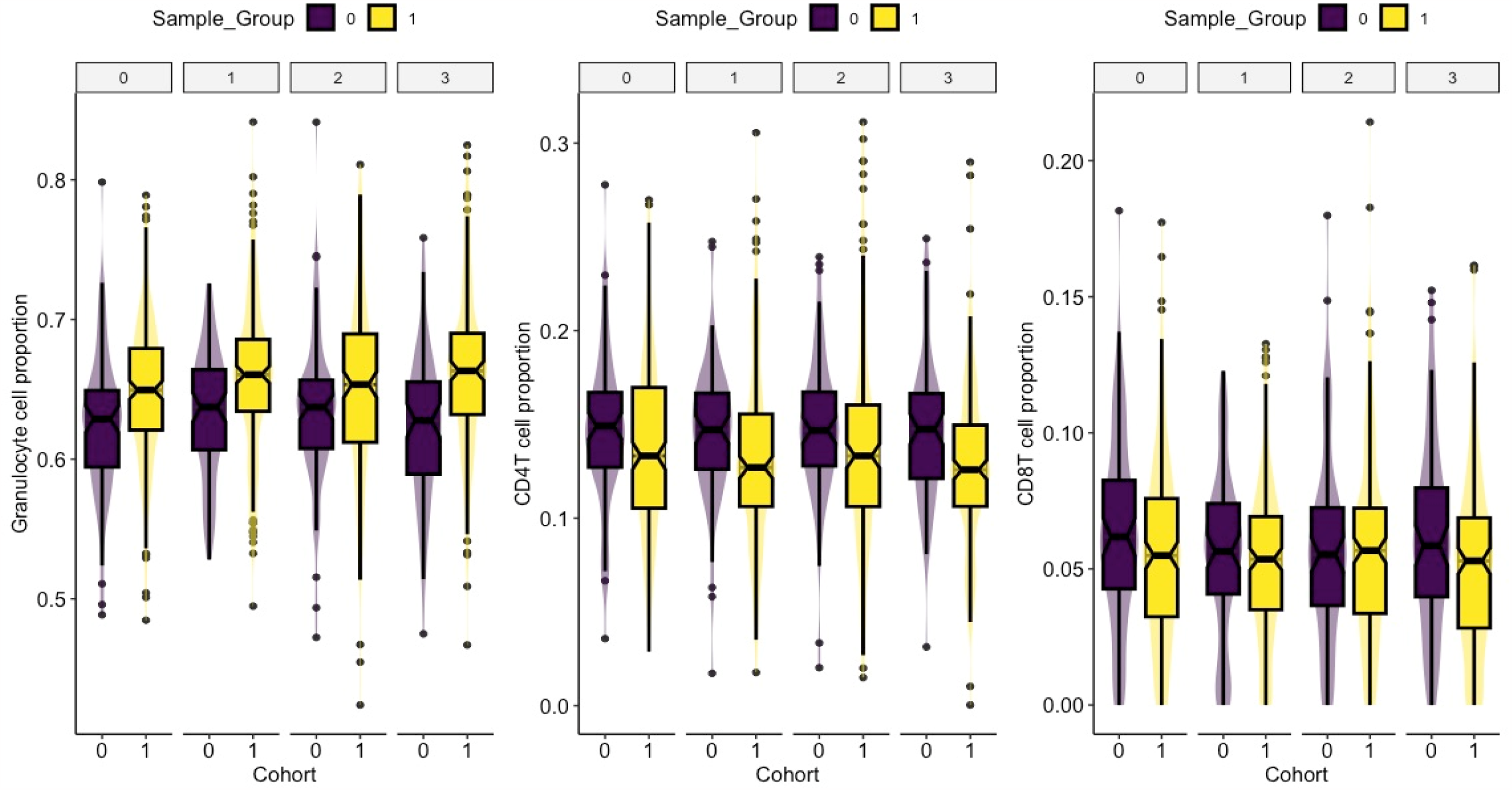
Blood cell composition in sporadic Parkinson Disease individuals.

### DNA Methylation differs between sPD and healthy controls

We conducted a cross-sectional analysis by comparing methylation profiles between sPD cases and HC at enrollment. This analysis revealed 81,604 differentially methylated sites between PD and HC. Applying a cutoff of a 2% absolute change in average methylation (Δβ), resulted in 5,183 retained positions. The top differentially methylated positions are presented in **Table 3**. PD individuals exhibited hypermethylation in 2,683 DMPs and hypomethylation in 2,495 DMPs (**Fig2A**). Approximately 30% of DMPs were enriched in promoter regions (**Fig2B**). Gene ontology analysis revealed that these DMPs were associated with genes involved in diverse cellular processes, including several with specific functions in the brain (“Focal adhesion”, “Cholinergic synapse”, “Glutamatergic synapse”, “Dopaminergic synapse”) (**Fig2C**).

**Figure 2.**
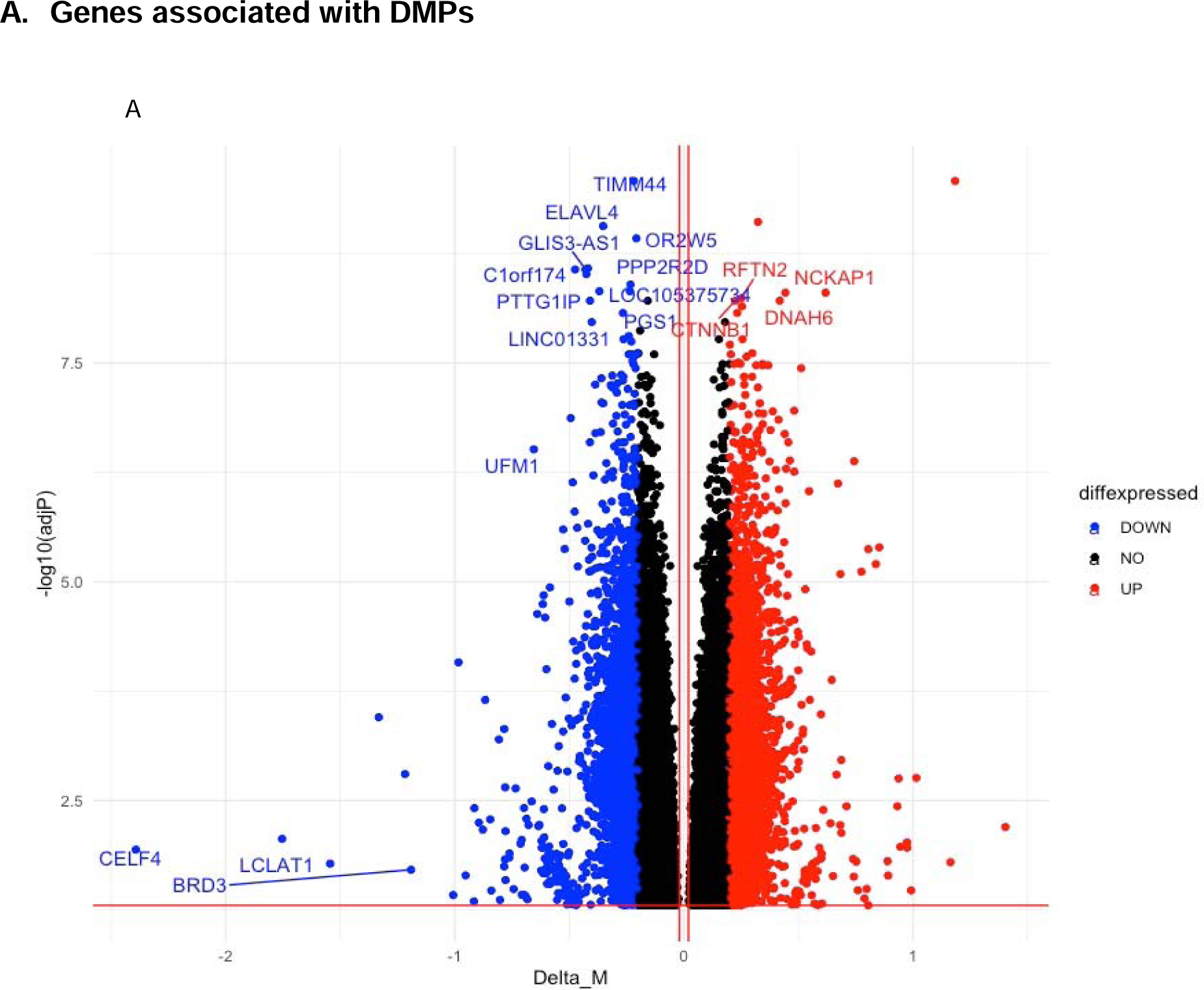
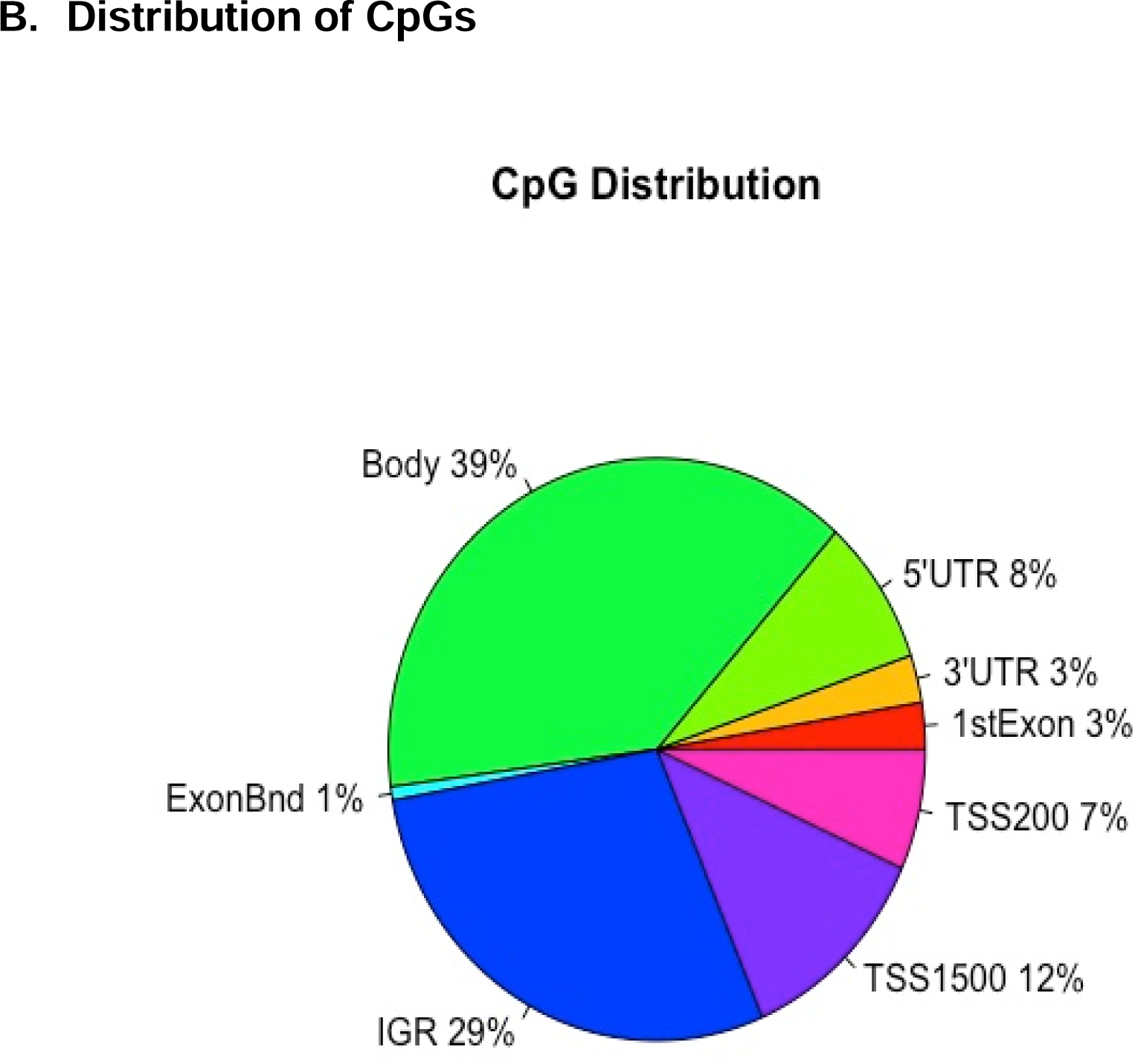
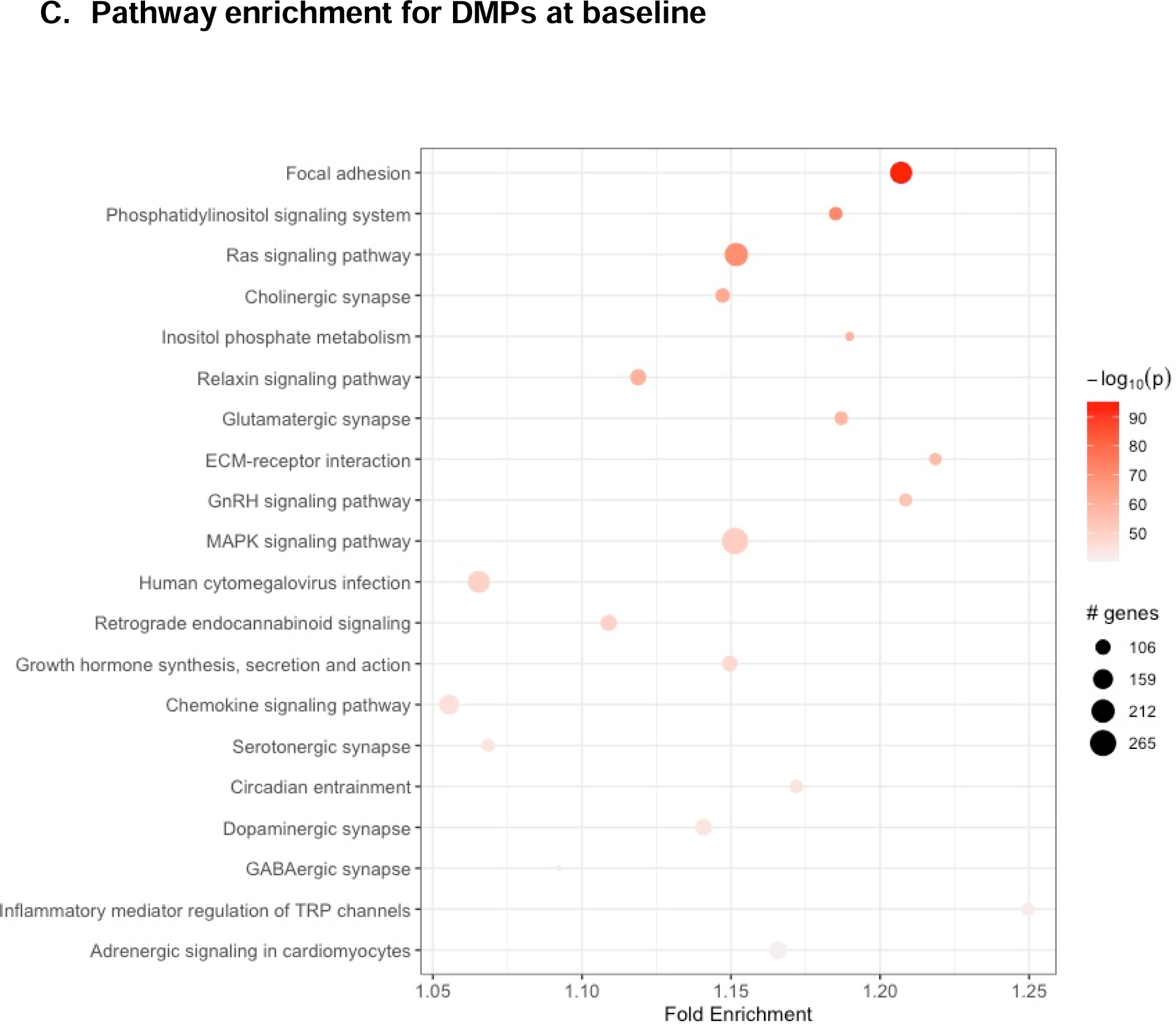
Differentially Methylated Probes (DMPs) Volcano plot of differentially methylated cytosine-guanine sites (CpGs). Each point represents an individual probe. Nearest associated gene is presented. UP=Hypermethylation; Down=Hypomethylation. Of note, differential methylation is presented as M values (log of B values).

**Table 3.** Top 20 differentially methylated probes (DMPs) at baseline.

| CpG | Chr | Position | Relation to TSS | Nearest associated gene | logFC | FDR |
| --- | --- | --- | --- | --- | --- | --- |
| cg18213661 | 11 | 93681423 | IGR |  | -0.2946842 | 0.00868945 |
| cg25403368 | 5 | 130554320 | IGR |  | -0.1673612 | 0.00035482 |
| cg06378142 | 19 | 50119633 | Body | *PRR12* | -0.1543515 | 0.03815787 |
| cg13388618 | 2 | 238814771 | Body | *RAMP1* | -0.1355725 | 0.00063473 |
| cg17509989 | 5 | 176798049 | Body | *RGS14* | -0.1284415 | 0.00682996 |
| cg15355235 | 18 | 35001518 | Body | *CELF4* | -0.1198882 | 0.01153713 |
| cg10326673 | 2 | 30669757 | TSS1500 | *LCLAT1* | -0.1107039 | 0.04497683 |
| cg03646189 | 5 | 2756101 | IGR |  | -0.109624 | 0.00989133 |
| cg06087988 | 2 | 46567260 | Body | *EPAS1* | -0.109153 | 2.34E-05 |
| cg14121685 | 15 | 56299462 | IGR |  | -0.1054255 | 1.80E-05 |
| cg27586797 | 5 | 13664584 | IGR |  | 0.19857441 | 0.0160684 |
| cg06612594 | 20 | 43682078 | Body | *STK4* | 0.133571 | 0.01565868 |
| cg01938825 | 7 | 1563708 | IGR |  | 0.13325065 | 0.04997444 |
| cg12858166 | 6 | 33033176 | 3'UTR | *HLA-DPA1* | 0.11387422 | 0.03364694 |
| cg18661872 | 5 | 131436611 | IGR |  | 0.11101836 | 4.20E-07 |
| cg05804568 | 5 | 159592696 | IGR |  | 0.1099409 | 2.63E-10 |
| cg07796016 | 1 | 152779584 | TSS1500 | *LCE1C* | 0.10961198 | 0.0417662 |
| cg26251192 | 14 | 74003199 | TSS1500 | *ACOT1* | 0.10695994 | 0.00160629 |
| cg24853868 | 1 | 146555624 | IGR |  | 0.10477105 | 0.00406238 |
| cg05194426 | 10 | 135343193 | Body | *CYP2E1* | 0.10295235 | 7.68E-06 |

Since methylation at one site can be dependent on the methylation status of nearby CpGs, we decided to interrogate DMRs. Consequently, 13,071 differentially methylated regions (DMRs), between sPD and HC at baseline, were identified. Using a logFC cutoff of ≥ ±0.05 to focus on positions with substantial differential methylation, we identified 5,281 DMRs. The top 20 DMRs are presented in **Table 4**, and gene ontology analysis similarly indicated a connection with brain-specific functions (“Dopaminergic synapse”, “Tyrosine metabolism”, “Notch signaling pathway”) (**Fig3**). A complete list of DMPs and DMRs results at each time point are presented in **Supplementary Tables 1-6**.

**Figure 3.**
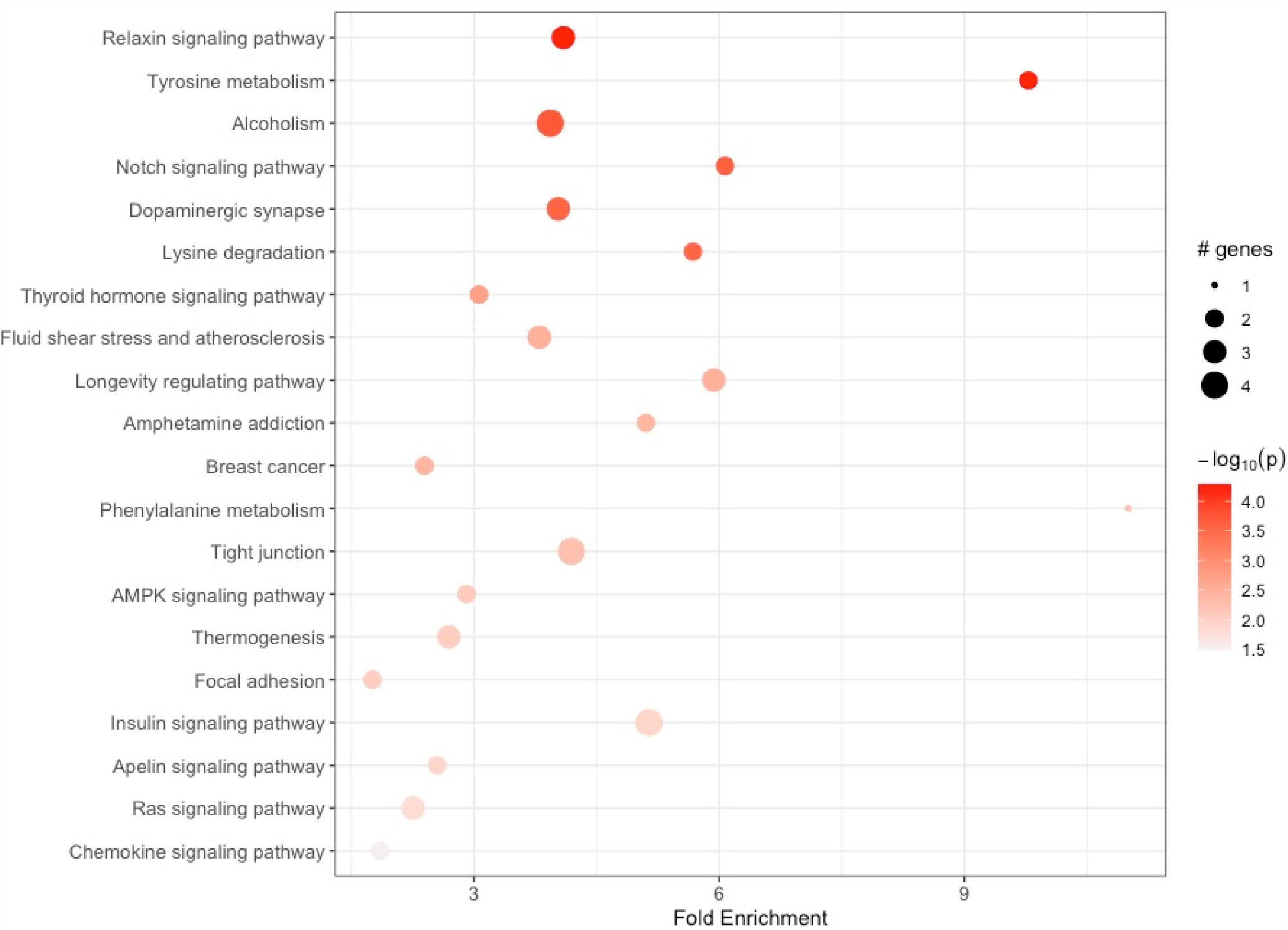
Pathway enrichment for Differentially Methylated Regions (DMRs) at baseline.

**Table 4.** Top 20 significant differentially methylated regions (DMRs) at baseline.

| Chromosome | Start | End | CpG | FDR | Max $\Delta\beta$ | Gene |
| --- | --- | --- | --- | --- | --- | --- |
| *Hypomethylated* |  |  |  |  |  |  |
| chr5 | 159592081 | 159592696 | 3 | 7.43E-10 | -0.1063226 | N/A |
| chr5 | 131436611 | 131436890 | 2 | 8.35E-07 | -0.1030687 | N/A |
| chr10 | 135340445 | 135343248 | 22 | 2.48E-05 | -0.0981492 | CYP2E1 |
| chr20 | 43681878 | 43682078 | 2 | 0.01865295 | -0.097719 | STK4 |
| chr11 | 1474588 | 1475899 | 7 | 2.87E-06 | -0.0899853 | BRSK2 |
| chr1 | 232007841 | 232008510 | 2 | 8.71E-05 | -0.0888528 | DISC1 |
| chr16 | 19777410 | 19778059 | 4 | 0.00648286 | -0.0842839 | IQCK |
| chr13 | 30053833 | 30055092 | 6 | 0.00095081 | -0.0756862 | MTUS2, |
| chr8 | 143751325 | 143751796 | 7 | 0.00430513 | -0.0740626 | PSCA,<br>JRK |
| chr5 | 179740743 | 179741120 | 4 | 0.04429012 | -0.0697626 | GFPT2 |
| *Hypermethylated* |  |  |  |  |  |  |
| chr2 | 46567260 | 46567451 | 2 | 4.66E-05 | 0.09948858 | EPAS1 |
| chr2 | 30669385 | 30670304 | 12 | 0.01696551 | 0.09649313 | LCLAT1 |
| chr15 | 56299462 | 56299636 | 2 | 1.40E-05 | 0.09613655 | N/A |
| chr12 | 7900072 | 7900651 | 3 | 0.00298332 | 0.09112004 | CLEC4C |
| chr5 | 176796807 | 176798049 | 7 | 0.00116872 | 0.0875691 | RGS14 |
| chr1 | 115825602 | 115825641 | 2 | 0.02418216 | 0.08476224 | N/A |
| chr11 | 1412270 | 1414627 | 9 | 0.00147298 | 0.08060391 | BRSK2 |
| chr1 | 101003634 | 101003924 | 3 | 0.02727681 | 0.07447863 | GPR88 |
| chr7 | 948246 | 949834 | 6 | 0.00280961 | 0.07007024 | ADAP1 |
| chr6 | 32305106 | 32305145 | 2 | 0.00142024 | 0.06978072 | C6orf10 |

### DNAm influences gene expression in sPD at baseline

To delve deeper into the relationship between DNAm changes and gene expression, we analyzed RNASeq data from PPMI. 75 significantly differentially expressed genes between sPD and HC were identified, with 71 being upregulated and 4 downregulated **(Supplementary Table 7)**. Integration of both differentially methylated sites and expressed genes showed 20 genes that were hypomethylated and overexpressed and one gene, *CTSH* (beta = 0.005; adjusted P value = 0.022), that was hypermethylated and associated with reduced expression (**Fig4**).

**Figure 4.**
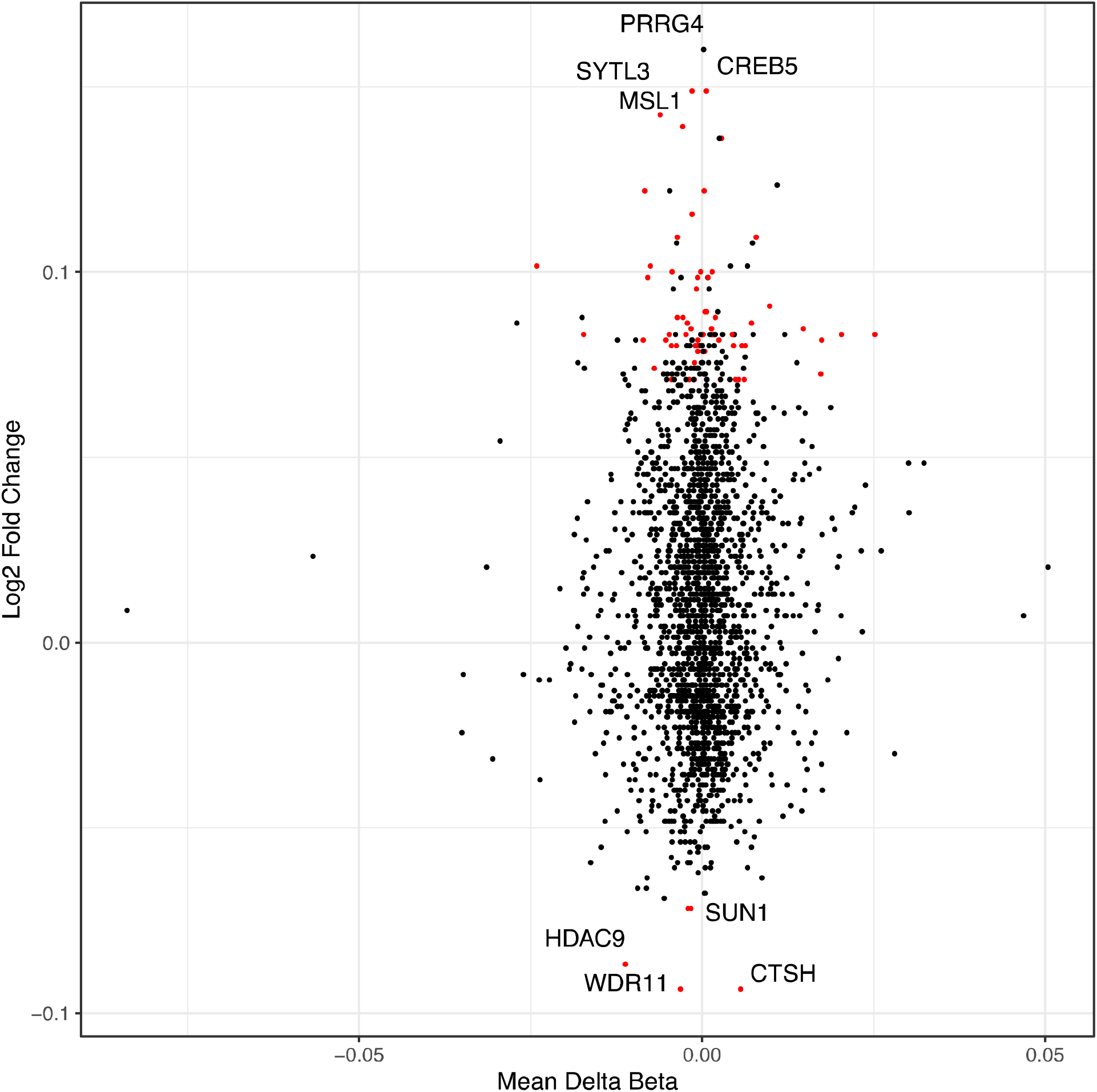
Overlap of differentially methylated and differentially expressed genes. Volcano plot of differentially methylated genes and differentially expressed genes. Each point represents an individual probe. Nearest associated gene is presented. UP=Hypermethylation; Down=Hypomethylation.

### Longitudinal Analysis

We extended our analysis to longitudinally profile samples collected over a span of 3 years. Demographic characteristics at each time point are detailed in **Table 2**. Linear regression models with cluster-robust standard errors identified 579 positions that significantly changed over time in sPD. To ascertain that age and LEDD were not the primary drivers of the longitudinal epigenetic alterations observed in sPD, we additionally assessed the relationship between methylation changes and these variables. We also found that several hypomethylated CpGs (7) were near the *CYP2E1* gene, while several hypermethylated CpGs (4) associated with the *NDRG4* gene (**Table 5**).

**Table 5.** Top Longitudinal changes in sPD cases.

| CpG_Site | Coefficient | SE | FDR | $\Delta\beta$ over three years | Nearest Gene |
| --- | --- | --- | --- | --- | --- |
| cg10596483 | 0.31088382 | 0.01116813 | 1.75E-115 | -0.0247078 | JRK |
| cg17416644 | 0.53757281 | 0.00913787 | 1.50E-273 | -0.0189683 | BRSK2 |
| cg21941354 | 0.2000633 | 0.00637213 | 7.49E-136 | -0.0170572 | PSCA |
| cg11445109 | 0.15685194 | 0.00880022 | 6.66E-59 | -0.0150986 | CYP2E1 |
| cg04008252 | 0.2226635 | 0.00607315 | 3.09E-165 | -0.0148353 | NPFFR2 |
| cg05473257 | 0.16573402 | 0.00645931 | 6.16E-103 | -0.0127294 | CYP2E1 |
| cg24984195 | 0.34316818 | 0.00607679 | 2.96E-263 | -0.0114182 | UBE2I |
| cg21299458 | 0.17011373 | 0.0043338 | 3.15E-179 | -0.0102815 | SCARF2 |
| cg19469447 | 0.1371077 | 0.00581347 | 5.06E-91 | -0.0100304 | CYP2E1 |
| cg03134882 | 0.19896345 | 0.00548764 | 5.23E-163 | -0.0100248 | CYP2E1 |
| cg17273911 | 0.21393103 | 0.00433243 | 1.45E-230 | -0.0092127 | N/A |
| cg09977703 | 0.19796984 | 0.00326851 | 5.65E-281 | -0.0089486 | PPP5C |
| cg05194426 | 0.25591425 | 0.00867698 | 5.36E-125 | -0.0089095 | CYP2E1 |
| cg01873886 | 0.25430321 | 0.00524984 | 4.87E-226 | -0.0089066 | BMP4 |
| cg23400446 | 0.14237323 | 0.00617015 | 4.17E-88 | -0.0087313 | CYP2E1 |
| cg10862468 | 0.19063746 | 0.0061232 | 2.34E-134 | -0.0083889 | CYP2E1 |
| cg12158483 | 0.18460978 | 0.00499163 | 5.51E-167 | -0.0083389 | BAIAP2L2 |
| cg12754571 | 0.51125879 | 0.00774553 | 2.83E-303 | -0.0076333 | LOC148824 |
| cg06378142 | 0.5259074 | 0.01546718 | 1.59E-150 | 0.03692832 | PRR12 |
| cg07504457 | 0.47968392 | 0.01020797 | 5.86E-219 | 0.02384914 | VPS28 |
| cg05725404 | 0.35295282 | 0.0064493 | 1.86E-255 | 0.02259907 | NDRG4 |
| cg00758881 | 0.24403566 | 0.00682108 | 2.27E-160 | 0.02230257 | NDRG4 |
| cg17457090 | 0.2751549 | 0.00549316 | 5.79E-234 | 0.02229852 | NDRG4 |
| cg27113419 | 0.27983815 | 0.00544638 | 4.67E-240 | 0.02203448 | NDRG4 |
| cg06211550 | 0.31122216 | 0.00835066 | 1.54E-168 | 0.02119219 | N/A |
| cg22772380 | 0.43071726 | 0.0069473 | 5.76E-287 | 0.01959139 | ZNF12 |

## Discussion

This study delves into the emerging role of epigenetic changes in PD and other neurodegenerative diseases. Specifically, we conducted a cross-sectional and longitudinal analysis of DNA methylation in whole blood samples from individuals diagnosed with sPD within the well-characterized and phenotyped PPMI cohort. This data also allowed us to examine DNAm patterns in a group of medication-free individuals at baseline, allowing us to uncover DNAm alterations more closely tied to disease mechanisms rather than medication-induced changes. Moreover, the longitudinal tracking of these individuals, coupled with comprehensive clinical and demographic data, offers a comprehensive view of PD’s DNAm dynamics. Significantly, despite our use of blood-derived data, many of these DMRs were linked to genes with established roles in brain function (e.g. *CTSH, NDRG4, CYP2E1, BRSK2*).

By comparing DNAm across a span of three years, we identified 579 DMPs, even after controlling for age and LEDD. These findings suggest a potential role of DNAm in disease progression. Of interest, several hypomethylated DMPs were associated with the *CYP2E1* gene that was also a highly significant DMR at baseline. While other groups have reported on differential methylation in this gene on cross-sectional data ^10, 24, 25^, our analysis demonstrates that this continues to be the case on follow up, suggesting a role of aberrant DNAm in *CYP2E1* in disease progression. Cytochrome P450 2E1, encoded by *CYP2E1*, plays a pivotal role in generating potentially toxic metabolites linked to dopaminergic degeneration. Furthermore, a SNP in this gene was identified as a genetic risk factor for PD in a Swedish cohort ^26^. *CYP2E1* polymorphisms have also been suggested as indicators of susceptibility to DNA damage in individuals exposed to pesticides, a finding of significance given the well-established association between pesticide exposure and PD ^27, 28^. Additionally, we also found several hypermethylated genes associated with the *NDRG4* gene; the protein encoded by this gene contributes to the maintenance of intracerebral BDNF levels and is required for cell cycle progression and survival in primary astrocytes. This gene has been found to have reduced expression in the substantia nigra and cinglulate gyrus of individuals with PD ^29, 30^. Future studies analyzing DNAm at longer intervals will be essential to gain a more comprehensive understanding of the link between DNAm and clinical progression, Most of the differentially methylated positions at each time point were situated within gene bodies. Gene body methylation refers to the methylation of CpG sites in the transcribed regions of genes. Growing evidence suggests that methylation in CpG-sparse regions, such as enhancers and gene bodies, can profoundly influence gene expression ^31^. While methylation in promoter regions typically silences genes, intragenic methylation may have a more a varied effect, such as promoting gene expression, gene silencing, or alternative splicing [35]. Therefore, further exploration through functional studies of these intragenic methylation sites may provide crucial insights into their role in PD.

To explore the link between DNAm and gene expression, we analyzed RNA Sequencing data in individuals with sPD. This analysis unveiled differences in gene expression profiles between sPD and HC individuals. Additionally, a comparison between differentially methylated genes and differentially expressed genes revealed an overlap of 21 genes. Here, we identified that the *CTSH* gene was hypermethylated and with reduced expression. CTSH encodes the Cathepsin H protein, a lysosomal cysteine protease involved in lysosomal protein metabolism ^32, 33^. Lysosomal dysfunction has been associated with PD ^34-36^, and lysosomal related genes are associated with increased PD risk. As such, abnormal DNAm of this protein emerges as a promising novel avenue for investigating PD pathogenesis. Additionally, while we were only able to perform a cross-sectional analysis of RNASeq data, origination and analysis of longitudinal gene expression data will aid in contextualizing these findings and their role in disease progression.

Our analysis of methylation profiles from whole blood also allowed us to investigate the abundance of specific blood cell types in sPD. At baseline, individuals with sPD exhibited an increase in granulocytes that persisted upon follow-up, in agreement with previous reports Our study also revealed a decrease in CD4T and CD8T cells upon follow-up (3 years after enrollment in PPMI). These changes may indicate immune system dysregulation in sPD, a finding of interest given the potential role of inflammation in the disease ^37, 38^.

These findings also suggest that DNAm could be explored as a marker for PD diagnosis and disease monitoring, likely as part of a broader biomarker panel. This will likely require the use of machine learning algorithms ^39^. Furthermore, as we move towards an era where the inclusion of pre-symptomatic individuals in clinical trials for PD becomes feasible, characterizing these changes in the prodromal stage of PD will be valuable for understanding the mechanisms underlying the disease onset and assessing the potential of DNAm as a biomarker for identifying individuals at high risk of progressing to motor symptoms.

Future research will also leverage the complete PPMI dataset, including genetic-PD individuals and validate our findings in other cohorts. This will be of importance given that most individuals included in PPMI are of European descent. As such, it is imperative to perform further analyses in diverse populations to ascertain if there are population specific methylation signatures that could explain PD risk. Additionally, there is a lack of easily accessible DNAm data in large PD cohorts. This study underscores the importance of collecting these biological data in future cohort studies, which will aid in replication and validation of this and subsequent epigenetic studies.

A limitation of our analysis is the absence of environmental data for these subjects. As mentioned earlier, DNAm can be influenced by environmental exposures. Future directions will involve analyses on environmental factors such as pesticide exposure and lifestyle factors like smoking and alcohol consumption collected as part of PPMI. Further functional characterization of some of these findings will be vital for uncovering new disease mechanisms and potential intervention avenues. Also, integration of DNAm and other biomarkers will be informative. As an example, now that PPMI has data on alpha-synuclein seed amplification assay (αSyn-SAA)^40^, this analysis will be rerun comparing individuals who are αSyn-SAA positive vs those who are αSyn-SAA negative.

In summary, our study provides evidence that alterations in the methylome in PD are discernible in blood, evolve over time, and reflect cellular processes linked to ongoing neurodegeneration. These findings lend support to the potential of blood DNA methylation as an epigenetic biomarker for PD. To fully comprehend DNA methylation changes throughout the progression of PD, additional profiling at longer intervals and during the prodromal stage will be necessary.

## Acknowledgments

PPMI, a public-private partnership, is funded by the Michael J. Fox Foundation for Parkinson’s Research and funding partners, including 4D Pharma, Abbvie, AcureX, Allergan, Amathus Therapeutics, Aligning Science Across Parkinson’s, AskBio, Avid Radiopharmaceuticals, BIAL, Biogen, Biohaven, BioLegend, BlueRock Therapeutics, Bristol-Myers Squibb, Calico Labs, Celgene, Cerevel Therapeutics, Coave Therapeutics, DaCapo Brainscience, Denali, Edmond J. Safra Foundation, Eil Lilly, GE HealthCare, Genentech, GSK, Golub Capital, Gain Therapeutics, Handl Therapeutics, Insitro, Janssen Neuroscience, Lundbeck, Merck, Meso Scale Discovery, Mission Therapeutics, Neurocrine Biosciences, Pfizer, Piramal, Prevail Therapeutics, Roche, Sanofi, Servier, Sun Pharma Advanced Research Company, Takeda, Teva, UCB, Vanqua Bio, Verily, Voyager Therapeutics, the Weston Family Foundation and Yumanity Therapeutics

## Author Contributions

PGL, DK, TS contributed to the conception and design of the study; PGL, BB, SD contributed to the acquisition and analysis of data; PGL, BB, SL, TS, DK contributed to drafting a significant portion of the text.

## Data Availability

The data that support the findings of this study are available from the corresponding author on reasonable request.

